# Large-Scale Analysis Reveals Racial Disparities in the Prevalence of ADHD and Conduct Disorders

**DOI:** 10.1101/2023.10.25.23297549

**Authors:** Noha Shalaby, Sourav Sengupta, Jamal B. Williams

## Abstract

**Objective:** The primary purpose of this study is to highlight trends in the prevalence of Attention Deficit/Hyperactivity Disorders (ADHD) and Conduct Disorders (CD) between non-Hispanic White and non-Hispanic Black populations and identify potential diagnostic disparities between these groups.

**Methods:** De-identified electronic health record data on the TriNetX platform of patients diagnosed with ADHD, CD, or both between January 2013 and May 2023 from 50 healthcare organizations in the US were used to investigate racial and sex disparities in the prevalence of ADHD and CD diagnoses.

**Results:** With a cohort of 849,281 ADHD patients and 157,597 CD patients, non-Hispanic Whites were ∼26% more likely to receive ADHD diagnosis and ∼61% less likely to be diagnosed with CD than non-Hispanic Blacks. The mean age of diagnosis of ADHD was over 8 years higher for White patients than for Black patients, with a disproportionately higher number of White patients diagnosed in adulthood, compared to a comparatively negligible number of Blacks diagnosed with ADHD in the same age group. Additionally, Black females were the cohort least likely to be diagnosed with ADHD, while White females were the cohort least likely to be diagnosed with CD.

**Conclusions:** Race disparities exist between Black and White populations, and sex disparities exist within each population. More information is needed to determine contributors to these differences, although implicit biases and systemic racism may be key contributing factors. Presenting evidence and increasing awareness of culturally relevant diagnoses can reduce unconscious bias and move toward more informed and objective psychiatric evaluations.

## Introduction

Attention-Deficit/Hyperactivity Disorder (ADHD) and Conduct Disorder (CD) are behavioral conditions that affect approximately 10% and 3% of children in the United States, respectively.^1,2^ However, the subjective nature of symptom assessment and the apparent overlap in behavioral manifestations between these disorders pose a significant risk for misdiagnosis.^3,4^ This issue is further complicated by the cultural context in which symptoms are evaluated, leading to potentially inappropriate categorization of behaviors.^5–7^

Both ADHD and CD have high societal and economic costs. ADHD persists into adulthood in about one-half to two-thirds of cases.^8–11^ Moreover, despite being characterized as a neurodevelopmental disorder in childhood, there is a recent recognition of adult ADHD cases.^11,12^ Current studies investigate whether ADHD in adults stems from missed childhood symptoms or comorbid mental health disorders rather than a distinct ‘late onset’ presentation.^10–13^ The long-term burden of ADHD lies in its comorbidity with other psychiatric disorders and increased rates of accidents, occupational failures, criminality, and addiction.^8^ As for CD, about 50% of individuals have a remission of symptoms in adulthood, while the rest frequently grow up to have a high risk of substance abuse, display criminal behaviors, or develop personality disorders.^2,14^

There is growing evidence that minority populations are less likely to be diagnosed with ADHD and less likely to take medication for ADHD compared to non-Hispanic White populations.^5,6,15^ Additionally, Black and Hispanic children are more likely to be diagnosed with CD than non-Hispanic White children.^5,6^ We hypothesize that non-Hispanic Black populations exhibit a lower likelihood of ADHD diagnosis, and this difference in diagnosis rates between Black and White populations may be linked to an overrepresentation of ODD and CD diagnosis in Black communities. There is also consensus that males are two to four times more likely to be diagnosed with neurodevelopmental disorders than females.^5,16^ Although this sex discrepancy may be related to the interplay between biological and societal factors, there is concern over diagnostic bias contributing to the underdiagnosis of females with neurodevelopmental disorders.^16^

In this study, we utilized large-scale data from de-identified electronic health records of patients diagnosed with ADHD and/or CD to identify the discrepancies in ADHD and CD diagnosis between non-Hispanic White and non-Hispanic Black populations. We analyzed the distribution of age of diagnosis and sex differences for each disorder.

## Methods

### Data Collection

This retrospective study was conducted using the TriNetX Research Network, which provides access to approximately 117 million anonymized patient electronic medical records from nearly 80 healthcare organizations across 4 countries. We first acquired data from patients diagnosed with ADHD (ICD-10 code F90), totaling 1,659,318 records. The second cohort acquired were patients with a CD diagnosis (ICD-10 code F91), with 422,625 patient records. These data were collected on June 5th, 2023. Our primary focus was on patient records within the US, consisting of approximately 96 million patients from 57 healthcare organizations. TriNetX, LLC, complies with the Health Insurance Portability and Accountability Act (HIPAA), and the study only uses de-identified records and is exempt from institutional review board approval.

### Study design

With the high prevalence and increased knowledge of ADHD worldwide, the American Psychiatric Association’s (APA) 2013 update to the Diagnostic and Statistical Manual of Mental Disorders (5th ed.; DSM–5)^17^ included revised criteria for the ADHD diagnosis.^18^ Plotting the annual incidence of ADHD and CD diagnoses in our cohort data from 2000 to 2022 (**Figure S1**) showed a steep increase in cases in recent years, particularly following 2013. Therefore, only patients with an initial diagnosis after 2013 were counted in each cohort. Using patient demographics, all patient records from outside the US or patients whose location was marked as “Unknown” were excluded. The focus was narrowed down to two groups of interest, a non-Hispanic White group, and a non-Hispanic Black/African American group. Each of these groups was further divided by sex into males and females.

ICD-10 codes were used to stratify each cohort by presentation, with some individuals falling under multiple presentations. The ADHD cohorts were divided into ADHD, predominantly inattentive type (F90.0), ADHD, predominantly hyperactive type (F90.1), ADHD, combined type (F90.2), Other ADHD (F90.8), and Unspecified ADHD (F90.9). In this study, Other and Unspecified ADHD were combined under the label Unspecified ADHD. The CD cohorts were divided into Oppositional Defiant Disorder (ODD, F91.3), Childhood-onset CD (F91.1), Adolescent-onset CD (F91.2), Other CD (F91.8), and Unspecified CD (F91.9). In this study, Other and Unspecified CD were combined under the label Unspecified CD.

Each patient’s age of diagnosis (AoD) was determined by subtracting the year of the first recorded incidence of the disorder from the year of birth. An AoD of 0 indicates that the birth year was missing from the patient’s record.

### Analysis and Statistics

Fisher’s exact tests were used to determine the significance of the differences in the prevalence of each disorder and each disorder’s presentations across race and sex. The reference population numbers for non-Hispanic White, non-Hispanic Black, and the total male and female populations were derived using queries on the TriNetX platform.

Student t-tests were used to determine the significance of the difference in the mean of the AoD between White and Black patients diagnosed with either ADHD or CD.

Linear regression was applied to examine the relationship between the ADHD AoD of Black and White patients and the CD AoD of Black and White patients. The coefficient of determination or correlation (R^2^) was calculated to measure the strength of the linear relationships.

Chi-squared tests were used to determine the degree of association between race and sex in the prevalence of ADHD or CD diagnoses in a population by creating two-way contingency tables combining the race and sex distribution of each disorder.

All data processing, statistical analyses, and figure generation were conducted in R.^19–22^

## Results

Amongst US patients on the TriNetX platform, 32,489,776 were non-Hispanic Whites, including 17,111,169 (52.7%) females. The non-Hispanic Black population accounted for 8,702,848 individuals, 4,654,839 (53.5%) females. In the White group, 708,004 individuals (2.18%) were diagnosed with ADHD. Out of those, 313,138 (44.2%) were females. Additionally, 110,160 (0.34%) were diagnosed with ODD or CD, and 36,300 (33%) were females. In the Black group, 141,277 (1.62%) were diagnosed with ADHD, including 51,323 (36.3%) females and 47,437 (0.55%) were diagnosed with ODD or CD, including 17,047 (35.9%) females. The incidences of each diagnosis and its associated presentations in each race and sex cohort were normalized by their population, then by the White cohort for racial analysis and the White male cohort for the sex analysis (**Figures S2 & S3; Table S1**).

In **Figure 1A**, apart from ADHD hyperactive type (ADHD-HT), ADHD diagnoses are significantly less prevalent in the Black population than in the White population. In the total ADHD cohort, a diagnosis in the Black population is 26% (OR, 0.74; 95% CI, 0.73-0.74; P<0.0001) less prevalent than in Whites. ADHD inattentive-type (ADHD-IT) is 55% (OR, 0.45; 95% CI, 0.45-0.46; P<0.0001) less prevalent in the Black population, while ADHD-HT is 0.03% (OR, 1.03; 95% CI, 1.01-1.05; P = 0.007) more prevalent in the Black population, although this result is not clinically significant. For ADHD combined-type (ADHD-CT) and unspecified ADHD, a diagnosis is, respectively, 13% (OR, 0.87; 95% CI, 0.86-0.88; P<0.0001) and 17% (OR, 0.83; 95% CI, 0.82-0.84; P<0.0001) less prevalent in the Black population than in the White population.

**Figure 1.**
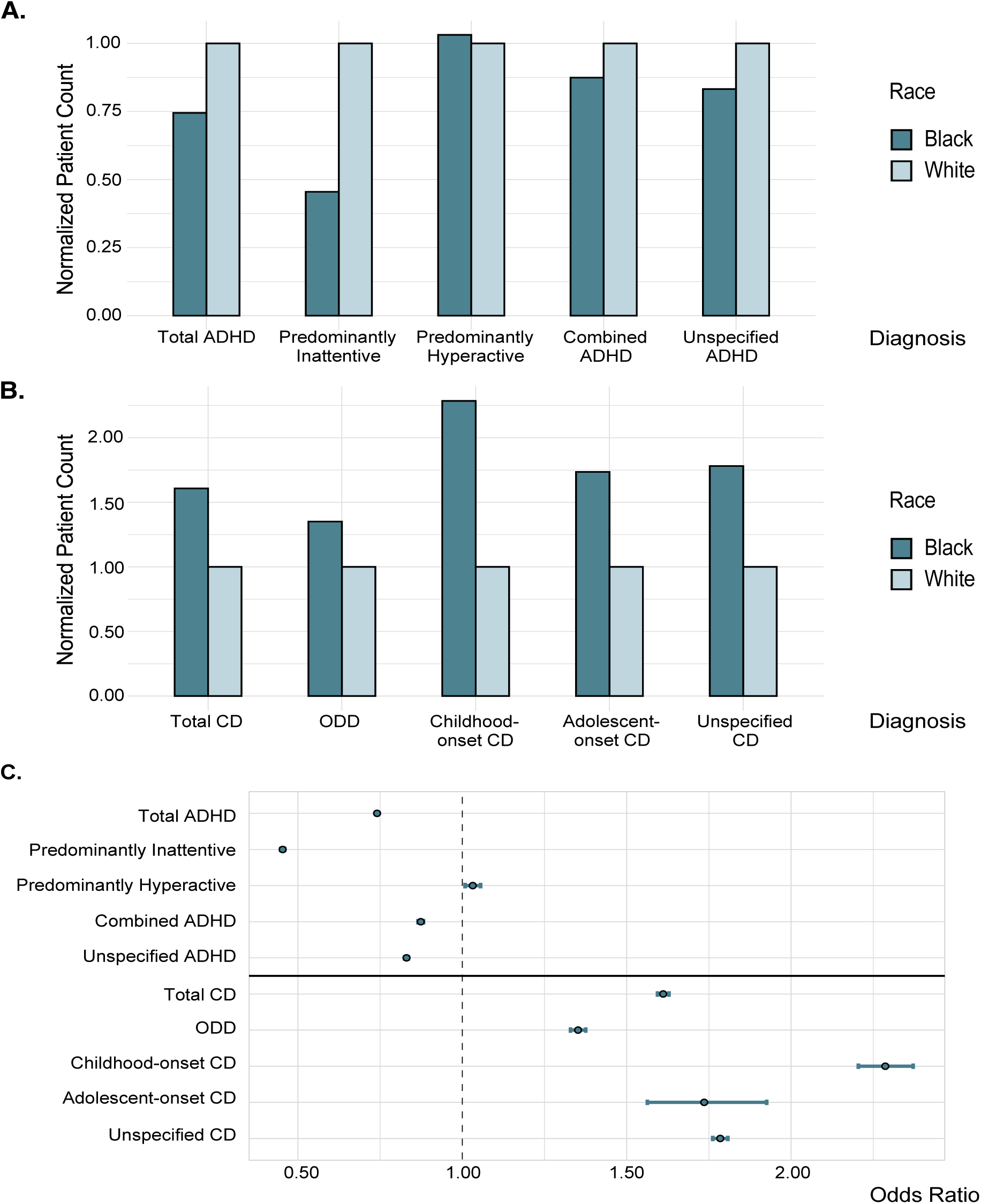
Prevalence of ADHD and CD Between Blacks and Whites. Normalized incidence of **A**. ADHD and its presentations, and **B.** CD and its presentations in Black and White populations. **C.** Odds Ratio and 95% confidence interval of the incidence of ADHD and CD and their presentations in the Black population compared to the White population. ADHD; Black N = 141,277, White N = 708,004; CD; Black N = 47,437, White N = 110,160.

In **Figure 1B**, CD diagnoses are more prevalent in the Black population than in the White population. In the total CD cohort, a diagnosis in the Black population is 61% (OR, 1.61; 95% CI, 1.59-1.63; P<0.0001) more prevalent than in the White population, while ODD is 35% (OR, 1.35; 95% CI, 1.33-1.37; P<0.0001) more prevalent. Childhood-onset CD and adolescent-onset CD are, respectively, 128% (OR, 2.28; 95% CI, 2.21-2.37; P<0.0001) and 73% (OR, 1.73; 95% CI, 1.56-1.92; P<0.0001) more prevalent in the Black population while unspecified CD was 78% (OR, 1.78; 95% CI, 1.76-1.81; P<0.0001) more prevalent than in the White population. **Figure 1C** shows the odds ratio and 95% confidence interval for the differences in the incidence for each disorder cohort and its presentations with the exact values listed in **Table 1**.

**Table 1:** Fisher’s Exact Test comparing the prevalence of ADHD and its presentations and CD and its presentations between the Black and White populations.

|  | Number of Black Patients | Number of White Patients | Odds Ratio | 95 % Confidence Interval | p-value |
| --- | --- | --- | --- | --- | --- |
| Total ADHD | 141,277 | 708,004 | 0.74 | 0.73-0.74 | <0.0001 |
| Predominantly Inattentive | 24,047 | 197,246 | 0.45 | 0.45-0.46 | <0.0001 |
| Predominantly Hyperactive | 9,416 | 34,073 | 1.03 | 1.01-1.05 | 0.007 |
| Combined ADHD | 53,895 | 230,083 | 0.87 | 0.86-0.88 | <0.0001 |
| Unspecified ADHD | 107,660 | 482,871 | 0.83 | 0.82-0.84 | <0.0001 |
| Total CD | 47,437 | 110,160 | 1.61 | 1.59-1.63 | <0.0001 |
| ODD | 17,826 | 49,248 | 1.35 | 1.33-1.37 | <0.0001 |
| Childhood-onset CD | 4,698 | 7,673 | 2.28 | 2.21-2.37 | <0.0001 |
| Adolescent-onset CD | 530 | 1,140 | 1.73 | 1.56-1.92 | <0.0001 |
| Unspecified CD | 34,590 | 72,490 | 1.78 | 1.76-1.81 | <0.0001 |

The AoD for each cohort was calculated, and we discovered that the average ADHD AoD of White patients is 23.9, which is significantly higher than the average ADHD AoD of Black patients at 15.7 (95% CI, 8.1-8.24; P < 0.0001), attributed to the relative increase in the diagnosis of Whites between the ages of 18 and 40 years old (**Figure 2A; Table S2**). Linear regression analysis resulted in a correlation coefficient of 0.86 between the AoD of ADHD in Black versus White patients (**Figure S4A**). However, a linear regression performed on a subset of patients 18 years old or younger yielded a higher correlation coefficient of 0.97 (**Figure S4B**). These data suggest that the AoD between Blacks and Whites with ADHD are essentially the same in childhood, yet there is a substantial increase in the diagnosis of ADHD in White adults compared to Blacks.

**Figure 2.**
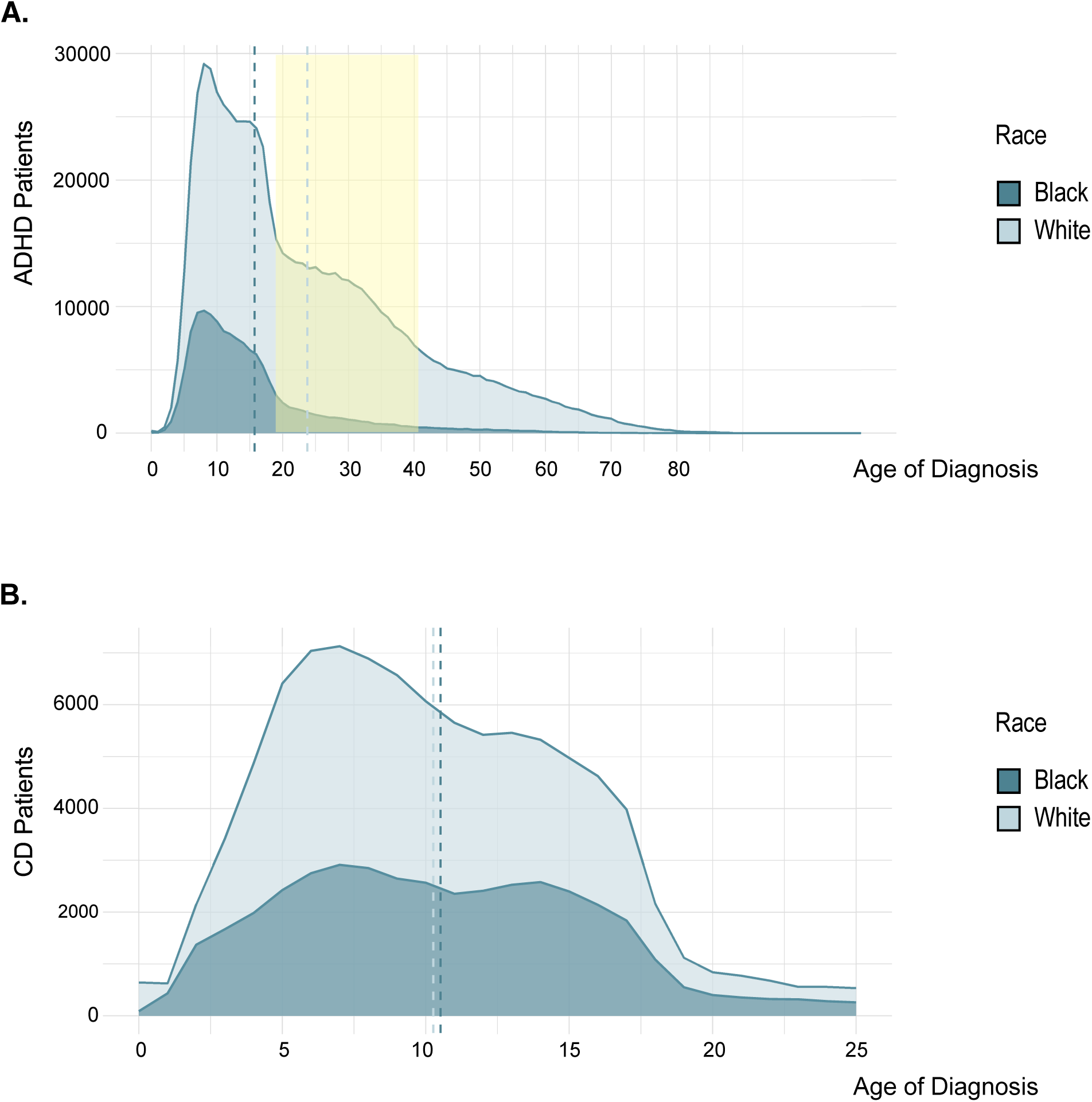
Age of Diagnosis Distributions. A. Density plot of the AoD distribution of ADHD in Black and White patients, **B.** Density plot of the AoD distribution of CD in Black and White patients.

The AoD distribution of Black and White CD patients highlights more parallel trends between the two groups (**Figure 2B**). Linear regression analysis shows a high correlation between AoD in CD cases in Black and White patients, with a coefficient of 0.97 (**Figure S4C**).

**Figure 3A** and **Table 2** show that Black females are the most underrepresented group across sex and race, being 59% (OR, 0.41; 95% CI, 0.41-0.42; P < 0.0001) less prevalent than White males. White females diagnosed with ADHD are 31% (OR, 0.69; 95% CI, 0.69-0.70; P < 0.0001) less prevalent, while Black males are 12% (OR, 0.88; 95% CI, 0.87-0.89; P < 0.0001) less prevalent. With ADHD-IT diagnosis in the Black male population is 47% (OR, 0.53; 95% CI, 0.52-0.54; P < 0.0001) less prevalent than the White male population, while diagnosis in the Black female population is 63% (OR, 0.37; 95% CI, 0.36-0.37; P < 0.0001) less prevalent. With ADHD-IT prevalence of diagnosis in Black males are 16% (OR, 1.16; 95% CI, 1.12-1.19; P < 0.0001) higher than in White males, while in Black females and White females, the prevalence of diagnosis is, respectively, 61% (OR, 0.39; 95% CI, 0.37-0.40; P < 0.0001) and 55% (OR, 0.45; 95% CI, 0.44-0.46; P < 0.0001) less. The prevalence of ADHD-CT diagnosis in Black females and White females are, respectively, 63% (OR, 0.37; 95% CI, 0.36-0.37; P < 0.0001) and 48% (OR, 0.52; 95% CI, 0.51-0.52; P < 0.0001) less than in White males. Unspecified ADHD diagnosis is 54% (OR, 0.46; 95% CI, 0.45-0.47; P < 0.0001) less in Black females, and 31% (OR, 0.69; 95% CI, 0.69-0.70; P < 0.0001) less in White females compared to White males.

**Figure 3.**
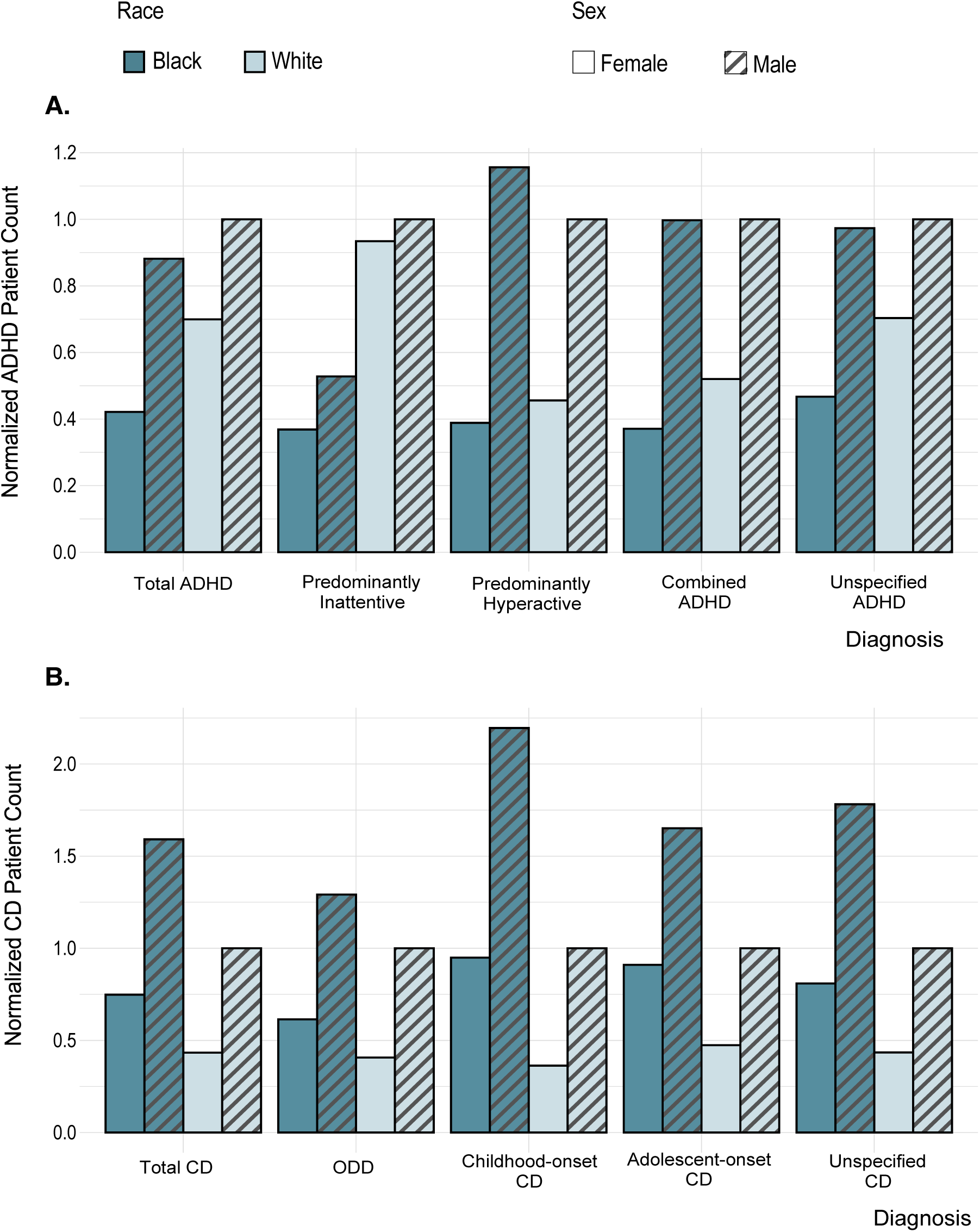
Sex Differences in ADHD and CD Prevalence Between Blacks and Whites. Normalized incidence of **A.** ADHD and its presentations and **B.** CD and its presentations in males and females in Black and White populations. ADHD; Black Female N = 51,323, Black Male N = 89,935, White Female N = 313,138, White Male N = 394,607; CD; Black Female N = 17,047, Black Male N = 30,381, White Female N = 36,300, White Male N = 73,837.

**Table 2:**
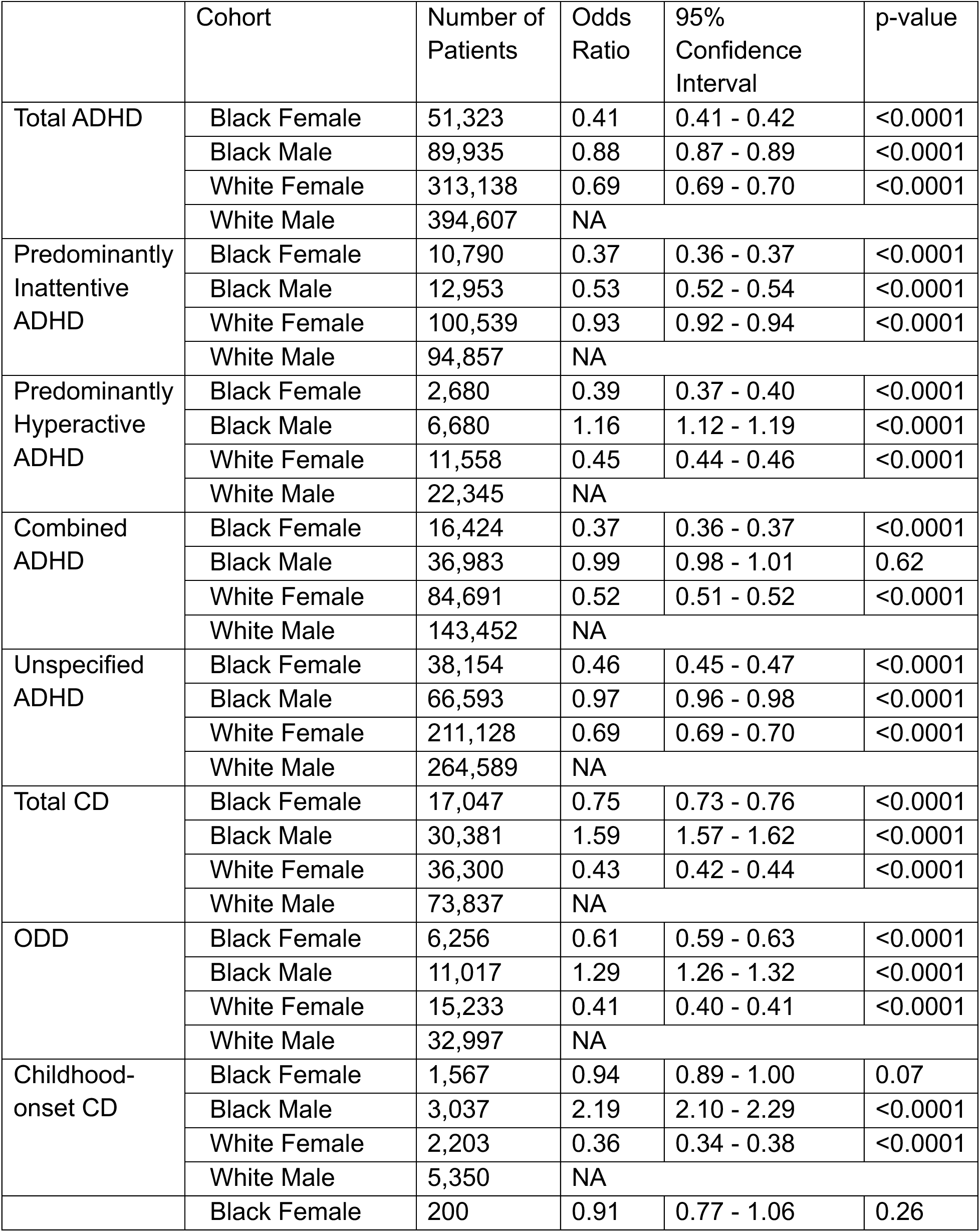
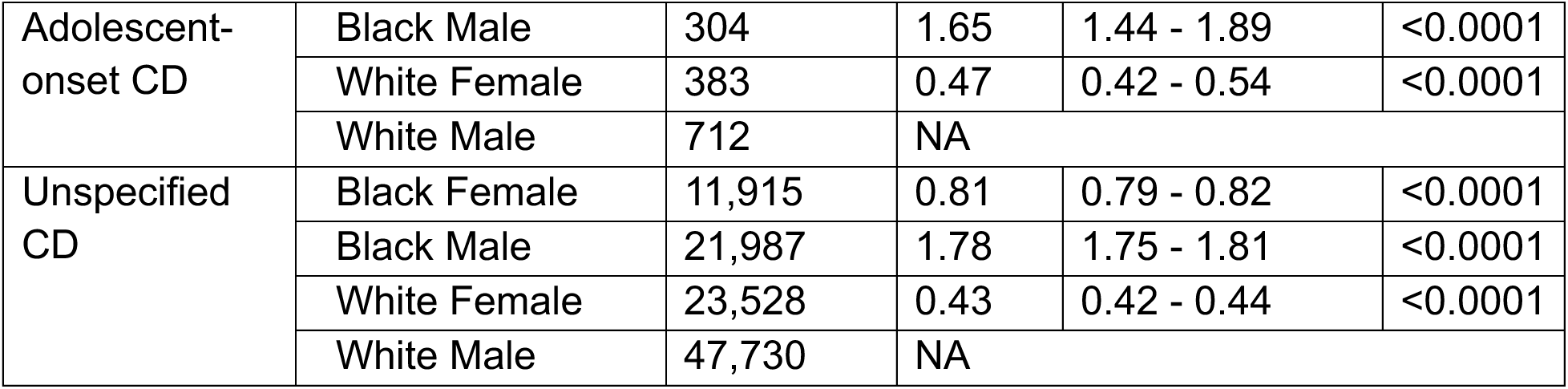
Fisher’s Exact Test comparing the prevalence of ADHD and its presentations, and CD and its presentations in each of the Black Female, Black Male and White Female populations to their prevalence in the White Male population.

**Figure 3B** and **Table 2** shows that compared to the White male population, total Black male CD diagnosis is 59% (OR, 1.59; 95% CI, 1.57-1.62; P < 0.0001) higher, while in Black females it is 25% (OR, 0.75; 95% CI, 0.73-0.76; P < 0.0001) less and in White females it is 57% (OR, 0.43; 95% CI, 0.42-0.44; P < 0.0001) less. ODD diagnosis in Black males is 29% (OR, 1.29; 95% CI, 1.26-1.32; P < 0.0001) more prevalent than in White males, but 39% (OR, 0.61; 95% CI, 0.59-0.63; P < 0.0001) less prevalent in Black females and 59% (OR, 0.41; 95% CI, 0.40-0.41; P < 0.0001) less prevalent in White females. Childhood-onset CD is 119% (OR, 2.19; 95% CI, 2.10-2.29; P < 0.0001) more prevalent in Black males and 64% (OR, 0.36; 95% CI, 0.34-0.38; P < 0.0001) less overall in White females than in White males. As for adolescent-onset CD, Black males have a 65% (OR, 1.65; 95% CI, 1.44-1.89; P < 0.0001) higher diagnosis prevalence, and White females have a 53% (OR, 0.47; 95% CI, 0.42-0.54; P < 0.0001) lower diagnosis prevalence than White males. Unspecified CD is 78% (OR, 1.78; 95% CI, 1.75-1.81; P < 0.0001) more prevalent in Black males, 19% (OR, 0.81; 95% CI, 0.79-0.82; P < 0.0001) less prevalent in Black females and 57% (OR, 0.43; 95% CI, 0.42-0.44; P < 0.0001) less prevalent in White females, all compared to prevalence in White males.

The odds ratio and 95% confidence interval for the prevalence differences in ADHD, CD, and their presentations of each race and sex cohort are plotted in **Figure S5** comparing **A.** Black male, **B.** White female, and **C**. Black female, all to White male. Pearson’s Chi-squared test results show a significant association between the categories of race and sex in each disorder **(Table S3)**.

## Discussion

ADHD is a complex neurodevelopmental disorder of varying presentations with social and cultural considerations. We hypothesized that Blacks are likely to have a lesser prevalence of an ADHD diagnosis and a higher prevalence of an ODD or CD diagnosis when compared to Whites. In this study, countrywide large-scale analysis supports our hypothesis, revealing that ADHD diagnoses are overrepresented in White patients compared to Blacks. Apart from ADHD-HT, all other presentations of ADHD have significantly less prevalence in Blacks than Whites. The most notable difference was found to be in the diagnosis of ADHD-IT. ADHD-IT is generally the most under-recognized and undertreated presentation of ADHD, with affected individuals having a low likelihood of receiving clinical and behavioral services.^23^ Our results indicate that this problem may be compounded in the Black population, with disproportionately lower diagnoses of Black patients with ADHD-IT potentially contributing to significant disparities in access to clinical and behavioral services and utilization.

Untreated ADHD-IT in adolescents and adults can be expected to impact an individual’s academic pursuits and career due to symptoms such as forgetfulness and difficulty maintaining attention for tasks, chores, or workplace responsibilities.^23^ In Black children, ADHD symptoms can be disproportionately misconstrued as willful or defiant behaviors, contributing to a greater likelihood of a diagnosis of ODD or CD and a corresponding lack or mismatch of appropriate interventions or inappropriate use of disciplinary strategies.^3,6,24^ A study using data from the 2011-2012 National Survey of Children’s Health found that White children were more likely to be diagnosed with ADHD alone, while Black children were more likely to be diagnosed with ADHD with an ODD or CD comorbidity.^25^ Our findings add to the characterization of this disparity, demonstrating a significantly higher prevalence of ODD and CD diagnoses in all presentations between Blacks and Whites. The cause of the gap in childhood-onset CD is unclear. Clinicians have demonstrated a reluctance to assign an early CD diagnosis for aggressive behavior in childhood, possibly expecting children to mature out of these patterns developmentally or, at times, giving an ODD diagnosis instead to avoid the stigma associated with a CD diagnosis.^2^ However, our results indicate that this cautionary approach may not be afforded to Black children as often.

Along with ADHD symptoms that persist from childhood into adulthood, there is a growing trend of adults presenting with inattention, disorganization, and impulsivity not recognized in childhood.^26–28^ There is skepticism about the diagnosis of ADHD in adulthood, as it is not fully understood and may be driven by individuals seeking stimulant medication with the symptoms more likely explained by other psychiatric or substancwe use disorders.^12,13,29^ Moreover, adult ADHD is not easy to identify. With the difficulty of obtaining a neuropsychiatric evaluation outside of childhood, especially for underprivileged populations, this job usually falls to primary care physicians (PCP) with little training in diagnosing complex psychiatric disorders.^26,27^ The high prevalence of unspecified types of ADHD and CD in our cohorts indicates that PCPs might diagnose neurodevelopmental disorders without a closer examination of the diagnostic criteria necessary to identify the disorder presentation or possible comorbid conditions.^26^

While most ADHD diagnoses take place before the age of 18 in both populations, our analysis reveals that adult ADHD is diagnosed much more prominently in White adults compared to Blacks. The Black population has a steady decline in ADHD diagnoses after adolescence. However, in White patients, there is a disproportionately higher number of patients diagnosed between 18 and 40 than in Black patients.

Receiving an ADHD diagnosis as an adult can be more complex, potentially requiring access to specialized or extended evaluations and necessitating greater expenditure of significant social capital. This disparity between the two racial groups could result from unequal access to general and psychiatric healthcare services in adulthood.

Furthermore, implicit biases stemming from societal or cultural differences could potentially lead to the misinterpretation of service-seeking behaviors as stimulant-seeking actions, thereby contributing to the underdiagnosis of Black adults who may have ADHD.^7,28^

Females are less likely to be diagnosed with neurodevelopmental disorders.^5,16,30^ Studies have shown that despite males displaying a higher prevalence of ADHD and CDs, females suffer from more severe symptoms, significant lifetime psychiatric comorbidities, and functional impairments.^5,16,30^ Apart from diagnostic bias, a potential contributing factor to sex disparities in these disorders is variation in symptom presentation. Females tend to exhibit more inattentive behavior and less hyperactivity, making them perceived as less disruptive than males, and their ADHD might go unnoticed or undiagnosed.^31,32^ As for CD, while males are inclined to display signs of proactive physical aggression, females are more likely to show relational, reactive aggression in bullying and manipulative behavior with less callous-unemotional traits.^30^ Beyond developmental gender differences, this could also suggest that females must exhibit more pronounced symptoms before being referred for or diagnosed with these disorders. Our analysis shows that Black females are the least prevalent group diagnosed with all presentations of ADHD. Apart from predominantly inattentive ADHD, White females are the second most underrepresented group in ADHD diagnoses. White females also appear to be the group least diagnosed with CDs.

Prior studies have revealed that conscious or unconscious bias can impact medical decisions and diagnoses.^6^ These studies indicate clinicians are more responsive to non-Hispanic White patients seeking an ADHD diagnosis and treatment, whereas Black students receive fewer referrals from schoolteachers and administrators.^5,6^ Moreover, obtaining a CD diagnosis will likely negatively impact a caregiver’s ability to detect inattentive or hyperactive behavior, limiting their access to psychiatric evaluations, medication, and therapy.^6^ It could also lead to harsher disciplinary measures and exclusionary practices in school that could further compound mental and behavioral challenges.^5,6^

One concern related to overdiagnosis and overtreatment of neurodevelopmental disorders like ADHD is the potential harm stemming from diverting resources from other populations who may be underdiagnosed or undertreated.^33^ Furthermore, the perception of ADHD as an overdiagnosed disorder in any racial group is likely.^34^

This study has some limitations. First, our lack of control subjects hindered our ability to perform more detailed analyses. Second, we did not have access to robust information about our patient cohorts’ socioeconomic status and insurance status, which is often a factor in a patient’s ability to receive clinical care, particularly mental healthcare and treatment.^5^ Third, with our reliance on ICD-10 codes, there were no symptom details to determine diagnostic accuracy. Fourth, we had very few quantitative measurements that could have been used to control confounding variables.

In conclusion, our analysis found race and sex disparities in ADHD and CD diagnosis in the US. The non-Hispanic Black population is less likely to be diagnosed with ADHD but more likely to be diagnosed with ODD or CD than non-Hispanic Whites. White patients get diagnosed with ADHD in adulthood more often than Black patients. Black females are the cohort least likely to be diagnosed with ADHD, and White females are the cohort least likely to be diagnosed with CD. Future work that integrates patients’ socioeconomic status and insurance status will give a deeper understanding of racial disparities. Detailed clinical symptom presentations, with quantifiable assessments compared to control cohorts, could be used to analyze further and measure accurate rates of over- and under-diagnosis in suspected populations. The disproportionally high rates of ODD and CD diagnoses carried by Black patients may indicate unconscious/implicit bias by healthcare practitioners and a corresponding tendency to miss underlying conditions that could better explain disruptive behaviors. Presenting evidence and increasing awareness of such disparities has effectively reduced unconscious bias and sustained the movement toward more culturally informed and objective psychiatric evaluations.^6^

## Data Availability

All data used in the present study is available through TriNetX at https://trinetx.com/

