## Supplemental Figures and Tables for "Large-Scale Analysis Reveals Racial Disparities in the Prevalence of ADHD and Conduct Disorders"

**Figure S1:** Line plot of the trend of **A**. ADHD diagnosis, and **B**. CD diagnosis between 2000 and 2022 in Black and White patients.

**A.**

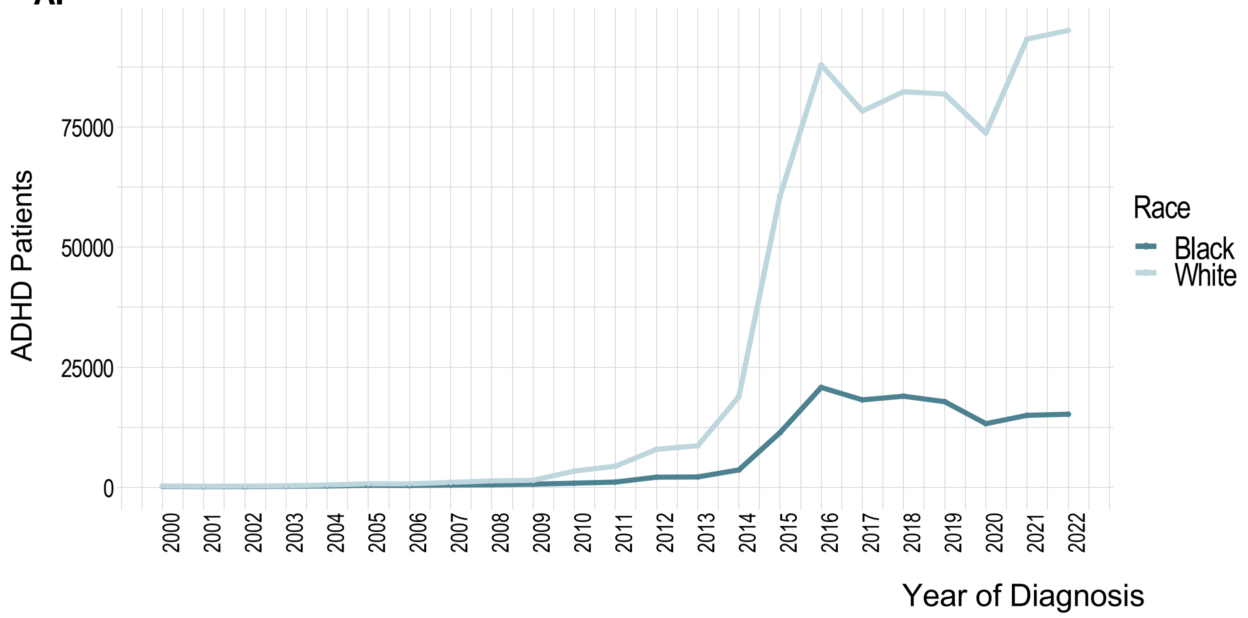


**B.**

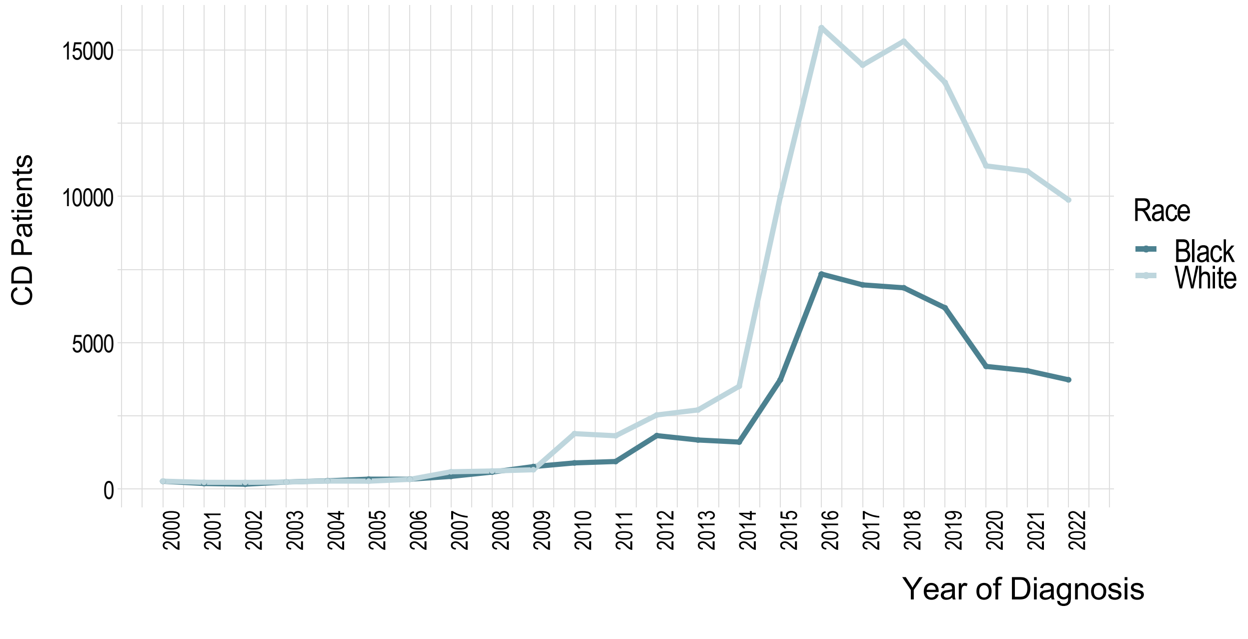

**Figure S2:** **A**. Incidence of ADHD and CD in Black and White populations. **B**. Normalized incidence of ADHD and CD in Black and White populations.

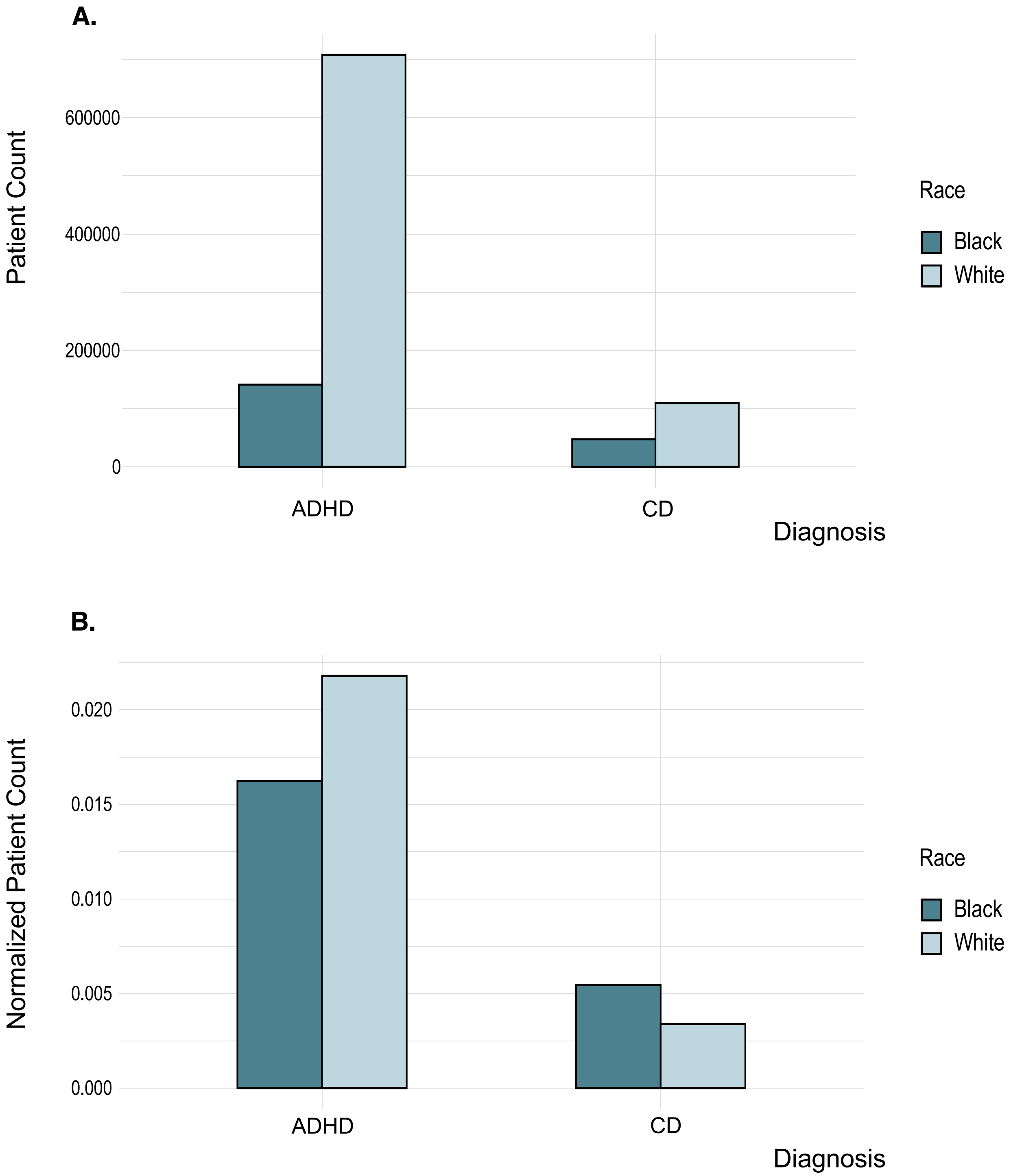

**Figure S3: A.** Incidence of ADHD, **B.** Normalized incidence of ADHD, **C.** Incidence of CD, and **D.** Normalized incidence of CD in Black Male, Black Female, White Male, and White Female populations.

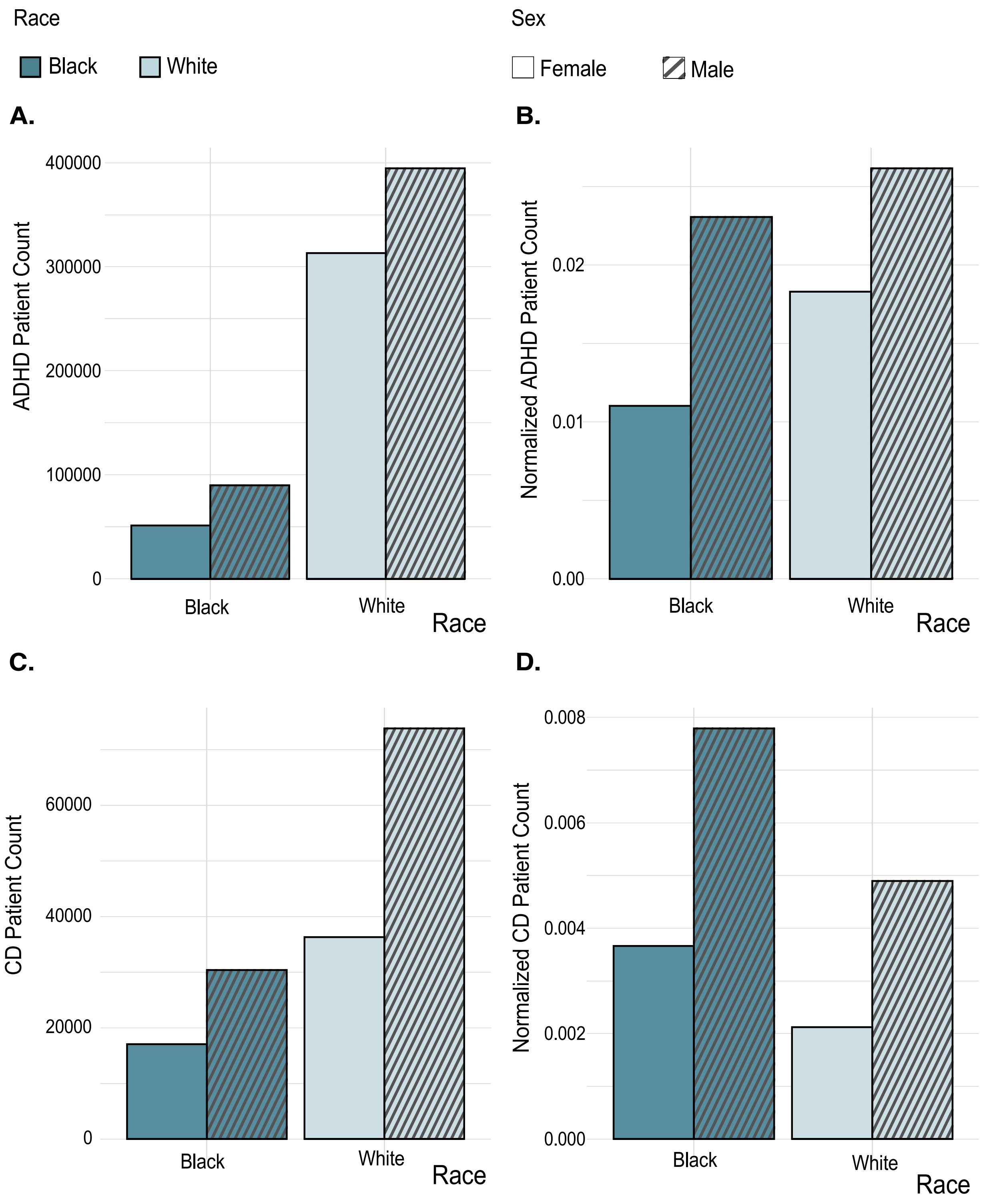

**Figure S4:** Linear Regression line plots show the linear correlation between Black and White patients; **A.** across all the ages of diagnosis of ADHD patients, **B.** in ADHD patients 18 years old and younger, and **C.** across all the ages of diagnosis of CD patients.

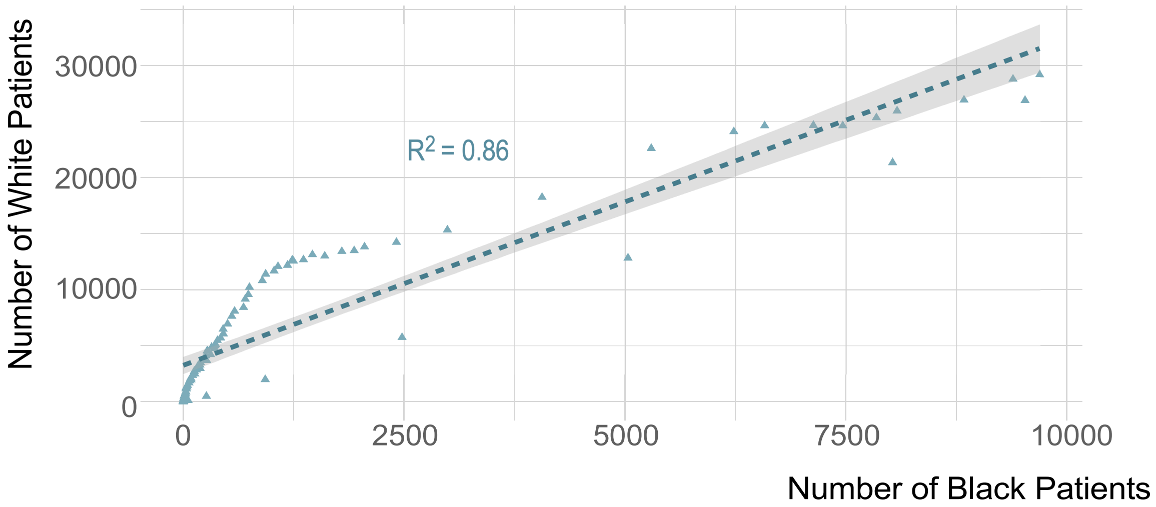


**B.**

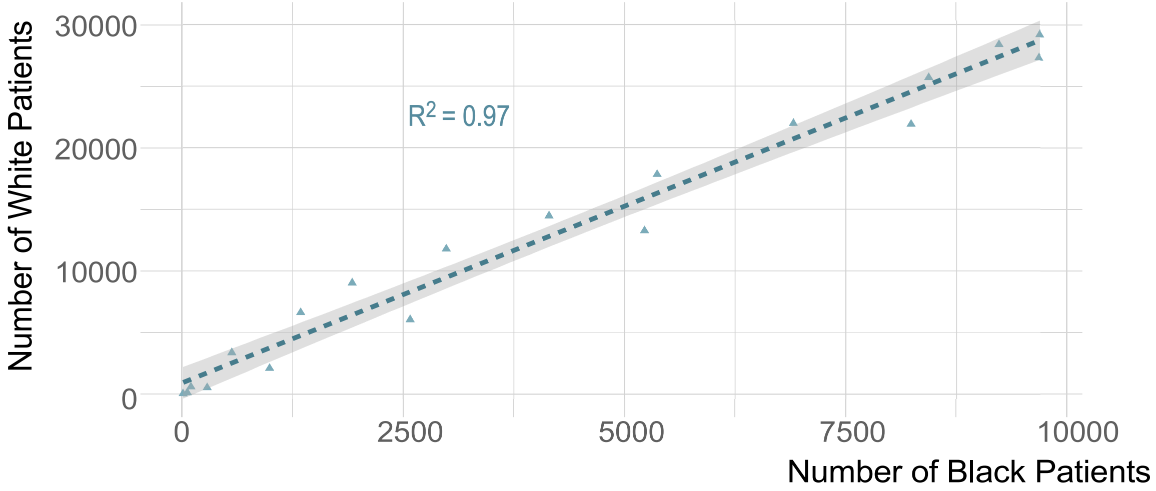


**C.**

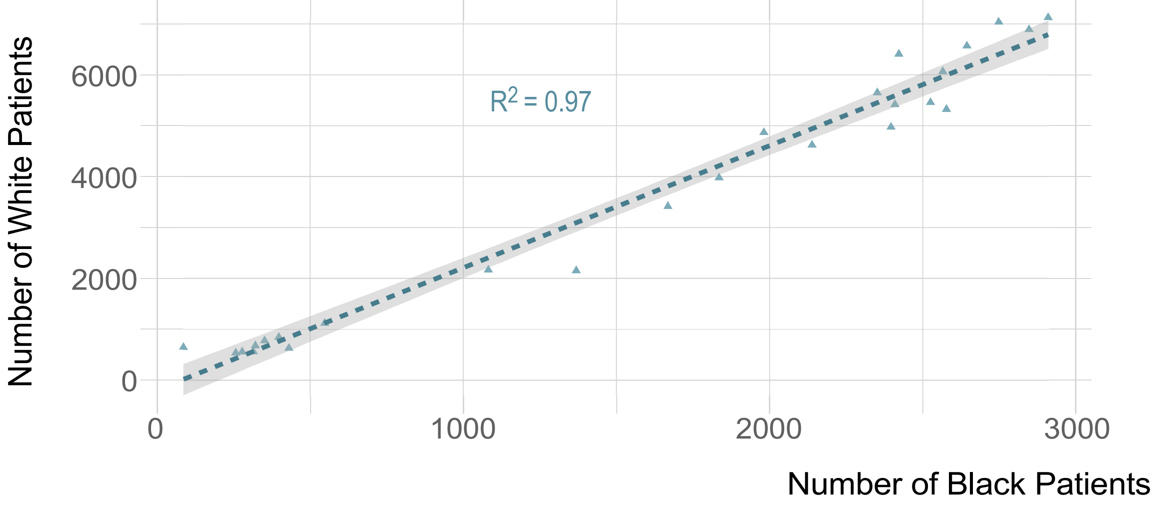

**Figure S5:** Odds Ratio and 95% confidence interval of the incidence of ADHD and CD and their presentations in **A.** the Black male population, **B.** the White female population, and **C.** the Black female population, all compared to the White male population.

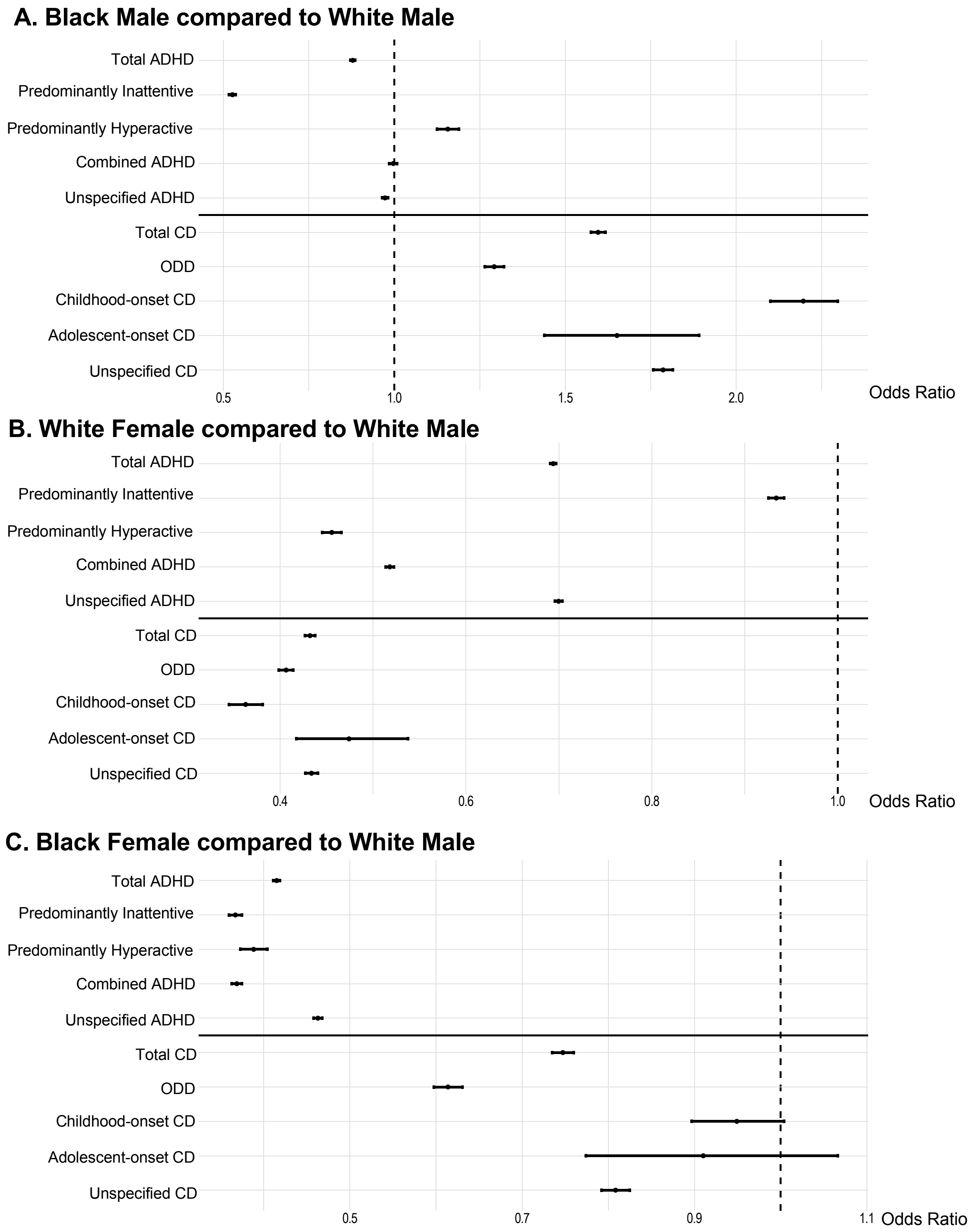

**Table S1:** Race and sex distribution of the patient population

| | Number of patients in group | % | Number of ADHD patients | % | Number of CD patients | % |
|---|---|---|---|---|---|---|
| Race | | | | | | |
| Black/African American | 8,702,848 | 21.1 | 141,277 | 1.62 | 47,437 | 0.55 |
| Sex | | | | | | |
| Female | 4,654,839 | 53.5 | 51,323 | 36.3 | 17,047 | 35.9 |
| Male | 3,899,810 | 44.8 | 89,935 | 63.7 | 30,381 | 64.0 |
| White | 32,489,776 | 78.9 | 708,004 | 2.18 | 110,160 | 0.34 |
| Sex | | | | | | |
| Female | 17,111,169 | 52.7 | 313,138 | 44.2 | 36,300 | 33.0 |
| Male | 15,083,418 | 46.4 | 394,607 | 55.7 | 73,837 | 67.0 |

**Table S2:** Statistical differences in the ages of first diagnosis of ADHD and CD between black and white patients

| | Black | White | $R^2$ | p-value |
|---|---|---|---|---|
| ADHD | | | | |
| Mean | 15.7 | 23.9 | 0.86 | <0.0001 |
| Range | 1 - 88 | 1 - 108 | | |
| ADHD (Children) | | | | |
| Mean | 9.00 | 9.51 | 0.97 | <0.0001 |
| Range | 1 - 18 | 1 - 18 | | |
| CD | | | | |
| Mean | 10.5 | 10.3 | 0.97 | <0.0001 |
| Range | 1 - 25 | 1 - 25 | | |

**Table S3:** ADHD and CD Pearson's Chi-squared test

| | | Black | White | $\chi^2$ | p-value |
|---|---|---|---|---|---|
| ADHD | | | | | |
| | Female | 51,323 | 313,138 | 3008.5 | <0.0001 |
| | Male | 89,935 | 394,607 | | |
| CD | | | | | |
| | Female | 17,047 | 36,300 | 131.81 | <0.0001 |
| | Male | 30,381 | 73,837 | | |
